# Longitudinal study of blood microbiome responses to milk reintroduction in healthy volunteers: A prospective, single-arm crossover study protocol

**DOI:** 10.1101/2025.05.16.25327766

**Authors:** Junhong Su, Bettina E Hansen, Zifan Wang, Zhongren Ma, Maikel P. Peppelenbosch

## Abstract

**Background:** The gut microbiome is crucial for maintaining overall host homeostasis and metabolism, but it can be significantly affected by dietary changes, leading to substantial temporal variations in microbial composition both within and between individuals. Despite these variations, fecal sampling remains the standard for microbiome assessment. Recently, the blood microbiome, which reflects the microbial DNA circulating in the bloodstream, has emerged as a potentially more stable and integrated alternative. Preliminary data suggest that analyzing blood microbiomes may provide more consistent insights than fecal samples.

**Method/Design:** This study will test the validity of the blood microbiome as a representative of the gut microbiome. Using a prospective, single-arm crossover trial, the researchers will evaluate the effect of milk consumption on the composition of milk-associated intestinal bacteria (MAB) in the blood of healthy volunteers aged 18-65. Participants will first abstain from dairy products, which is expected to reduce MAB cell-free DNA in the bloodstream. They will then reintroduce milk, during which MAB DNA is anticipated to reappear in the blood. Changes in blood MAB levels will be tracked using shotgun sequencing. Longitudinal sampling of this process will provide critical evidence for the existence of a common blood microbiome and refine the bioinformatic pipeline for its analysis.

**Conclusion:** This pilot trial will determine whether blood MAB is a valid surrogate marker for gut MAB, potentially offering a more consistent alternative to traditional microbiome assessments.

**Clinical Trial Registration:** NCT06944002

## Introduction

Ecological DNA (eDNA) is a concept that is gaining popularity for making comprehensive descriptions of all organisms present in an ecosystem. Gut microbiomes can be considered a specific variant of eDNA [1, 2]. A description of the gut microbiome should theoretically contain all different kinds of microorganisms in our gastrointestinal tract, including bacteria, viruses, archaea, fungi and other organisms (e.g. worms and phages). Accurately describing the gut microbiome is challenging due to spurious temporal variability and technical challenges (e.g. the commonly used 16S rRNA approach ignores many organisms) [3]. Despite these challenges, making accurate descriptions is essential as the gut microbiome plays critical roles in maintaining host homeostasis and health [4, 5].

To overcome the challenges in sampling the intestinal microbiome, most investigators currently rely on probing feces. We have pioneered the field in developing improved technical approaches; for example, we previously performed a double-balloon endoscopic study in healthy volunteers to characterize this microbiome in its entirety [6]. However, fecal samples have limitations: they poorly capture adherent (mucosal) flora, which interacts directly with epithelial cells and may influence their function more significantly than bacteria in the lumen [7, 8]. Additionally, fecal microbiomes are highly heterogeneous, with variations depending on which part of the sample is analyzed. Technological hurdles also exist, such as the resistance of many eukaryotes to DNA extraction protocols. Furthermore, diet greatly influences fecal microbiota composition, with meals like breakfast introducing different microbial communities compared to dinner [9]. These issues highlight the need for alternative methods that can provide stable, informative microbiome data predictive of host health.

An emerging approach involves analyzing the blood microbiome—a new method that integrates microbial signals from various body sites, particularly the gut [10]. The blood microbiome refers to the collection of cell-free DNA (cfDNA) fragments derived from microorganisms, rather than living organisms themselves. The mechanisms by which microbial DNA enters circulation are not fully understood [11], but the gastrointestinal microbiome is thought to be a significant source, especially when gut barrier integrity is compromised, which is often considered a common feature in chronic diseases. Blood-based microbiome signatures have shown promise as biomarkers for conditions such as diabetes, cardiovascular disease, and cancer, and they may even help distinguish between different cancer types [10, 12]. However, the field faces controversy due to bioinformatics challenges and potential artifacts [13, 14], underscoring the need for rigorous proof-of-concept studies. Our current research seeks to address this gap.

Building on previous findings, we observed that *Lactococcus lactis*, a milk-associated intestinal bacterium (MAB), is undetectable in human stool four days after stopping dairy intake. Upon reintroducing dairy, *Lactococcus lactis* quickly reappears [15, 16]. Although the precise half-life of bacterial DNA in blood remains unknown, evidence from cancer- and fetal-derived cfDNA suggests it is generally less than 24 hours [17]. Therefore, a 10-day dairy withdrawal should substantially reduce *Lactococcus lactis* cfDNA in the blood, with its reemergence following milk reintroduction.

In this paper, we present the rationale, design, methods, and analytical plan for a single-arm crossover trial that aims to test the effect of milk consumption on the blood microbiome in healthy volunteers. The specific aims are to examine: 1) MAB DNA counts in blood and fecal samples; and 2) other microbiome DNA counts (bacteria, viruses and Archaea) in these samples. We hypothesize that milk intake, known to significantly influence the fecal microbiome, will lead to detectable changes in blood DNA communities, reflecting a shared microbiome signature between blood and gut. Capturing this phenomenon through such a dietary interventional trial in volunteers will provide critical proof-of-concept for the blood microbiome technology we have developed.

## 2. Methods

### 2.1. Study design

We will perform a within-subjects crossover trial with a single arm to study the effect of milk on the blood and fecal Lactococcus levels in healthy volunteers. We shall first withdraw volunteers from dairy product consumption for 10 days. From previous work, we know that after four days of dairy product withdrawal, Lactococcus lactis is no longer present in the feces. As we expect half-life for bacterial cfDNA to be shorter than 24 hours, ten days of withdrawal should be sufficient to observe a substantial reduction in Lactococcus DNA counts (i.e. DNA fragments that originally came from a Lactococcus lactis bacterium) in the blood. Following the reintroduction of milk, we anticipate that DNA fragments from intestinal Lactococcus lactis will reappear in the circulation, though we do not yet know the exact kinetics involved.

This study is approved by the ethics committee of Erasmus Medical University Center Rotterdam in May 2025 (NL88008.078.24) and is registered at www.clinicaltrials.gov (NCT06944002), hence prior to participant’s enrollment. This study will be conducted in accordance with the principles of the Declaration of Helsinki (75th WMA General Assembly, Helsinki, Finland, October 2024) and the Medical Research Involving Human Subjects Act (WMO). Interested volunteers will be provided with detailed information about the study and will give written informed consent prior to participation. The volunteers will be studied multiple times (within-person crossover design) (**Figure 1**): a baseline sample of blood and stool will be collected before the onset of any dairy product abstinence. Ten days later, stool and blood samples will be collected again to verify that Lactococcus has indeed disappeared from the fecal and the blood DNA. Subsequently, volunteers will be instructed to drink 700 ml (an amount recommended by the national dietary guidelines of the United States [18]) during breakfast for 7 consecutive days. We have no information on the kinetics by which bacterial DNA will translocate to the blood; thus, we have chosen three time points to capture these dynamics: 24 hours, 48 hours, and one week after the reintroduction of dairy products. Volunteers will be free at this time to consume other dairy products as well during the study.

**Figure 1.**
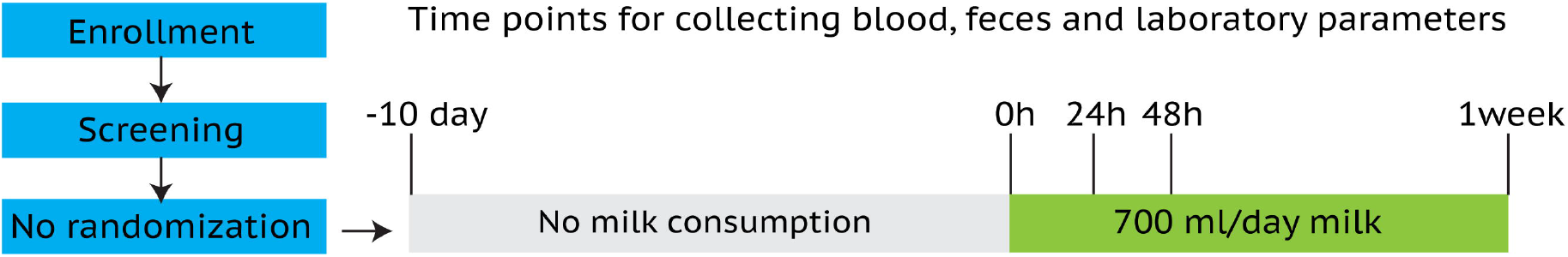
Study design. In this within-person crossover trial, the volunteers will be studied multiple times: a baseline sample of blood and stool will be collected before the withdrawal of any dairy product. Ten days later, the same samples will be collected again to verify if Lactococcus disappears from the stool and the blood DNA. Subsequently, volunteers will be instructed to drink 700 ml during breakfast for 7 consecutive days. Three time points will be chosen to capture the dynamics of Lactococcus DNA gut-to-blood translocation: 24 hours, 48 hours, and one week after the reintroduction of dairy products. The dairy products used in this study will be provided by the investigational team.

### 2.2. Participant recruitment

Healthy volunteers who are interested in participating in the study will be invited. To do this, flyers with contact information (for the secretary) will be posted within the university. All participants expressing interest will receive a study description document that they can read and evaluate at home. Participants will be given one week to make their decision. If they have additional questions, they may consult the leading investigator. One week later, participants will be approached again.

After providing sufficient information and answering any questions, participants who agree to participate will be requested by the authorized person to sign the informed consent form and to send it to the coordinating investigator or bring the form to the hospital to sign in the presence of the coordinating investigator. The process of the study is shown in **Figure 2**.

**Figure 2.**
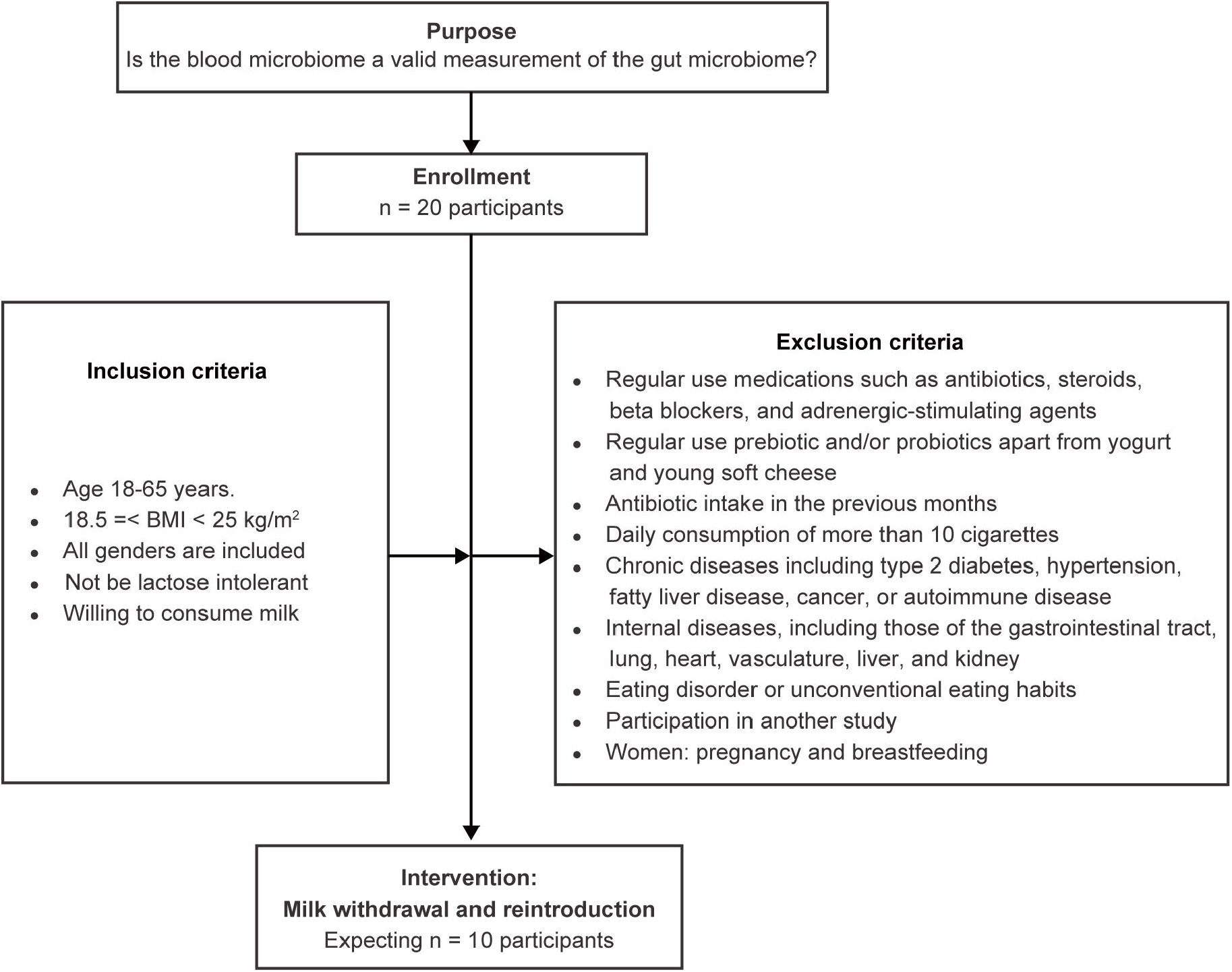
Study schema. This study requires to include data from at least 10 participants.

### 2.3. Inclusion criteria

1. Age 18-65 years
2. 18.5 =< BMI < 25 kg/m2
3. All genders included
4. Not be lactose intolerant
5. Willing to consume milk

### 2.4. Exclusion criteria (self-report)

1. Regular use medications such as antibiotics, steroids, beta blockers, and adrenergic-stimulating agents
2. Regular use prebiotic and/or probiotics apart from yogurt and young soft cheese
3. Antibiotic intake in the previous months
4. Daily consumption of more than 10 cigarettes, a minimum quantity known to significantly affect the gut microbiome [19]
5. Chronic diseases including type 2 diabetes, hypertension, fatty liver disease, cancer, or autoimmune disease
6. Internal diseases, including those of the gastrointestinal tract, lung, heart, vasculature, liver, and kidney
7. Eating disorder or unconventional eating habits
8. Participation in another study
9. Women: pregnancy and breastfeeding

### 2.5. Objective

#### 2.5.1. Primary objective

We aim to explore Lactococcus DNA counts in feces and blood before and after milk consumption. Lactococcus lactis only resides in the human intestine following milk consumption and is thus a modifiable part of the intestinal microbiome. The presence of Lactococcus DNA remnants in the blood will provide definitive evidence regarding the extent to which the blood microbiome captures any relevant changes in the intestinal microbiome.

#### 2.5.2. Secondary objective(s)

To explore other microbiome DNA counts (bacteria, viruses and Archaea) in faeces and blood before and after milk consumption.

#### 2.5.3. long-term objective

The final long-term objective is to develop technology that reliably provides descriptions of the dynamics in the microbiomes living in and on human organisms. The present study will show to what extent this is possible. Levels of non-milk-dependent organisms should remain relatively stable, whereas more milk-dependent organisms (e.g. Lactococcus and Bifidobacteria species) are expected to change. Demonstrating or not showing such changes will be instrumental in determining the usefulness of the blood microbiome technology for creating comprehensive, integrative descriptions of microbiomes residing on human hosts.

### 2.6. Power analysis

This is a pilot study. This research has never been conducted before, making it difficult to determine a definite sample size. Based on published studies showing significant change in the relative abundance of gut microbiome at the phylum level upon consumption of fermented milk in 6 volunteers (Firmicutes and Bacteroidetes; p<0.05 when comparing milk consumption vs. baseline) [20], and that the relative abundance of Bifidobacterium species increased following the consumption of human milk oligosaccharides by 7 volunteers over 7 days (p<0.05 when comparing four type of milk concentrations with baseline) [21], we have chosen to include 10 volunteers for this study. If this is not sufficient to convincingly demonstrate trends of Lactococcus lactis DNA during 7 days of milk consumption intervention, then blood microbiome studies may not be powerful enough to show meaningful changes in intestinal microbiome composition.

### 2.7. Study procedures (Table 1)

#### 2.7.1. Screening

A medical history and physical exam will be performed, including measurement of height, weight, blood pressure and BMI. Medical history will include the history of diseases, medication and smoking.

**Table 1.**
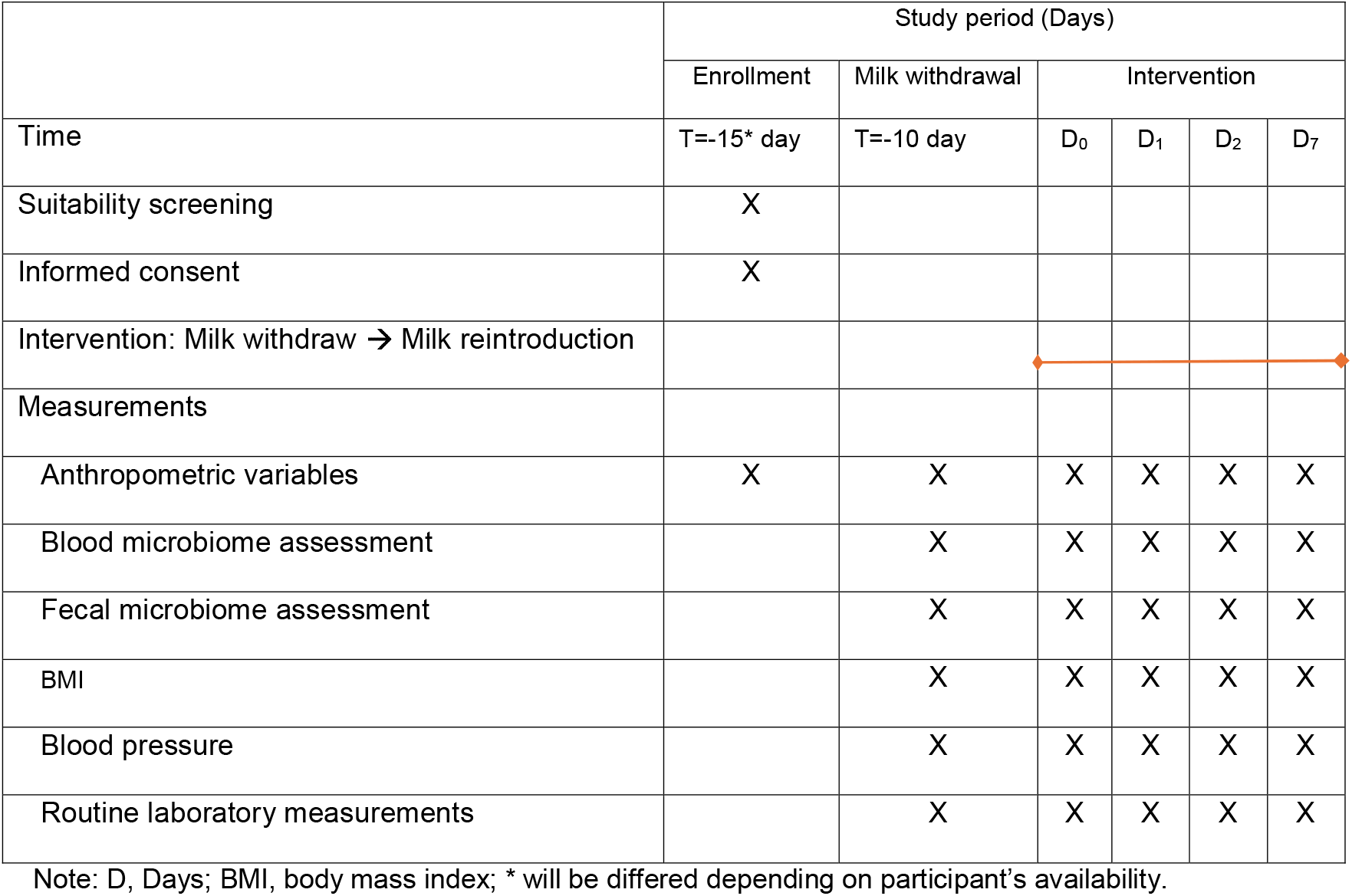
Overview of study procedures.

|  | Study period (Days) |  |  |  |  |  |
| --- | --- | --- | --- | --- | --- | --- |
|  | Enrollment | Milk withdrawal | Intervention |  |  |  |
| Time | T=-15* day | T=-10 day | D<sub>0</sub> | D<sub>1</sub> | D<sub>2</sub> | D<sub>7</sub> |
| Suitability screening | X |  |  |  |  |  |
| Informed consent | X |  |  |  |  |  |
| Intervention: Milk withdraw → Milk reintroduction |  |  |  |  |  |  |
| Measurements |  |  |  |  |  |  |
| Anthropometric variables | X | X | X | X | X | X |
| Blood microbiome assessment |  | X | X | X | X | X |
| Fecal microbiome assessment |  | X | X | X | X | X |
| BMI |  | X | X | X | X | X |
| Blood pressure |  | X | X | X | X | X |
| Routine laboratory measurements |  | X | X | X | X | X |

Note: D, Days; BMI, body mass index; \* will be differed depending on participant's availability.

#### 2.7.2. Baseline visit for milk withdrawal

In two weeks after screening visit, participants will be invited to visit the hospital in the morning for baseline visit for medical history, body composition (weight and BMI), blood pressure, and blood draw for blood microbiome. Participants will be requested to provide their fecal samples as well **(Table 2**). From this day, participants will withdraw all dairy products from diet life for consecutive 10 days.

**Table 2.** Primary, secondary and exploratory outcomes.

| **Outcomes** | **Brief description** | **Time frame** |
| --- | --- | --- |
| Primary |  |  |
| Blood MAB | Shotgun sequencing of whole blood DNA | -10, 0, 2, 3 and 7 days |
| Secondary |  |  |
| Other blood microbes | Shotgun sequencing of whole blood DNA | -10, 0, 2, 3 and 7 days |
| The gut microbiome | Shotgun sequencing of fecal DNA | -10, 0, 2, 3 and 7 days |
| Exploratory |  |  |
| BMI | Weight in kilograms and height in meters will be combined to report BMI in kg/m<sup>2</sup>. | -10, 0, 2, 3 and 7 days |
| Blood pressure | Systolic and diastolic blood pressure will be assessed with a blood pressure monitor. | -10, 0, 2, 3 and 7 days |

Note: MAB, milk-associated intestinal bacteria.

#### 2.7.3. Pre-intervention visit

In the morning of the eleventh day, participants will visit hospital to provide all information and samples as collected at baseline visit for milk withdrawal. After the completion of sample collection, participants will begin drinking milk during their breakfast for seven consecutive days.

#### 2.7.4 Intervention visit

On day 12 (24 hours after the time for milk reintroduction), day 13 (48 hours after the time of milk reintroduction) and day 18 (7 days after milk consumption), participants will visit hospital in the early morning at these time points, respectively. Participants will provide the same information and samples as collected during the baseline visit. There is no follow-up visit.

### 2.8 Microbiome analysis pipeline

#### 2.8.1 DNA extraction and shotgun sequencing

Fasting blood will be drawn by venipuncture by a certified phlebotomist at each time point. A total of 5 ml of whole blood with anticoagulant additive will be collected; 2.5 ml will be used for DNA extraction, and remainder will be used for plasma collection. Both sample types will be stored at - 80°C until further processing. The blood Lactococcus and other microbial DNA will be assessed by sequencing following the steps below: 1) Blood DNA will be extracted by a modified cetyltrimethylammonium bromide (CTAB) method [22]. Briefly, 1000 μl of CTAB lysis buffer at 65°C with lysozyme will be added to 250 μl of the whole blood and gently inverted. After centrifugation at 12 000 rpm for 10 min, the supernatant will be transferred into a 2.0 ml tube containing phenol/chloroform/isoamyl alcohol (25:24:1). Following another centrifugation, the supernatant will be moved into a new tube, mixed with 24:1 isoamyl alcohol and centrifugated again. The supernatant will be mixed with isopropanol, incubated at −20°C, and discarded. The precipitate will be washed with 1000 μl of 75% ethanol twice, dried and dissolved in dd H_2_O. Finally, 1 μl of RNase A will be added, and the mixture will be incubated at 37°C for 15 min; 2) DNA degradation will be monitored on 1% agarose gels, and concentration will be measured using Qubit® dsDNA Assay Kit in Qubit® 2.0 Flurometer (Life Technologies, CA, USA). Samples with an OD value between 1.8∼2.0 and contents above 1ug will be used for library construction; 3) Sequencing libraries will be created using NEBNext® Ultra DNA Library Prep Kit for Illumina (NEB, USA). First, index codes will be added to sequences. Then, DNA sample will be fragmented into approximately 350 bp by sonication, then end-polished, A-tailed, and ligated with adaptors for Illumina sequencing via PCR. Lastly, PCR products will be purified (AMPure XP system), with libraries analyzed for size distribution by Agilent2100 Bioanalyzer and quantified using real-time PCR; and 4) The clustering of index-coded samples will be performed on a cBot Cluster system. After cluster generation, DNA library will be sequenced on an Illumina PE150 platform, generating paired-end reads.

#### 2.8.3 Microbiome characterization

##### 1. Blood Lactococcus and other microbial DNA raw data processing

Illumina sequencing raw reads will be processed using Readfq to remove poor quality bases (threshold ≤ 38 bp), reads with excessive N bases, and significant adaptor overlaps. Host reads will be filtered using Bowtie2.2.4 software with parameters: --end-to-end, --sensitive, -I 200, -X 400 [23, 24].

##### 2. Metagenome assembly

Clean reads will be assembled into Scaffolds using MEGAHIT software with -presets meta-large, creating Scaftigs by removing N connections [25, 26]. Reads are compared to Scaffolds using Bowtie software, and fragments shorter than 500 bp are filtered for further analysis [27, 28].

##### 3. Gene prediction and abundance calculation

Scaftigs (≥ 500 bp) will be analysed for open reading frames (ORFs) using MetaGeneMark software, filtering out those shorter than 100 nt [29, 30]. CD-HIT software [31, 32] is applied to remove redundancy, creating a unique gene catalogue. Reads mapping to gene catalogue is conducted using Bowtie. Genes mapping to ≤ 2 reads are filtered out, and the remaining genes are used to calculate abundancy using a previously described method [28].

##### 4. Taxonomy prediction

Unigenes will be taxonomically identified using the Kraken database [33], followed by verification against the NCBI NR database [34]. The LCA algorithm in MEGAN [35] ensures meaningful species annotation, with taxonomy classifications used to create a table summarizing gene counts and abundance across taxa levels, including blood Lactococcus and other microbes.

#### 2.8.3 Microbiome taxonomic analysis

Krona tool [36] will be used to analyse the relative abundance of taxa in each sample and to create heatmap of abundance cluster. For microbiome composition analysis, PCA and NMDS (R vegan package) will be applied. ANOSIM will estimate the significance of changes in microbiome position. Taxonomic difference between groups will be assessed using Metastat and LEfSe software, with permutation test conducted and adjusted using the Benjamini–Hochberg false discovery rate. A P-value of less than 0.05 will be considered statistically significant. It should be noted that the presence and the abundance of the Lactococcus will be analysed separately from the pooled shotgun dataset.

#### 2.8.4 Assessment of the gut Lactococcus DNA and other microbial DNA

Fecal samples will be collected from each participant at each time point. Faecal DNA will be isolated as previously described [37]. The analysis of the faecal *Lactococcus DNA and other microbial DNA* will be performed based on the same procedures described above.

### 2.9. Statistical analysis

The change of relative abundance of blood Lactococcus DNA is the primary study parameter, which will be compared using repeated measurement ANOVA tests to determine whether there are statistically significant differences among five dependent samples. In a dependent sample, the same participants are measured five times at five different time points. Secondary study parameters will include the measurement of all other non-human organisms except Lactococcus. Repeated measurement ANOVA tests will also be used to evaluate whether there are statistically significant differences among five dependent samples.

## 3.0. Discussion

The concept of a common core blood microbiome in humans remains highly controversial across studies [10, 13]. Most of them are cross-sectional and lack appropriate control in addressing environmental contamination, which can pose a significant challenge in blood microbiome research and impede advancement in the field. Another major issue raised from these studies is the absence of prospective within-person controls for identifying, tracking and correcting possible false positive or negative outcomes resulting from high individual variation and shortcomings in bioinformatic pipeline [14]. In addition, while the gut microbiome is proposed to be the primary source of the blood microbiome, this hypothesis has yet to be proven. To address these barriers, a longitudinal study utilizing a nonharmful intervention known to effectively modify the gut microbiome may present a novel opportunity.

Milk is generally considered as safe and non-harmful, and it can significantly alter the gut microbiome, particularly by increasing beneficial bacteria, such as Lactococcus and Lactobacillus [38]. Given the gut-to-blood transition of gut microbiome-derived metabolites, it is reasonable to expect that gut microbiome-deprived microbial DNA, particularly those from milk-sensitive bacteria, may also be transferred into blood circulation, contributing the formation of the core blood microbiome. Considering these advantages, as well as the healthy and nutritional acceptance of milk, milk will be utilized as a gut microbiome modulator to explore MAB DNA in circulation. By examining the presence of milk associated Lactococcus DNA in both blood and fecal samples before and after milk reintroduction, we expect to demonstrate the responsiveness of the blood microbiome to dietary changes. This research has the potential to validate our blood microbiome technology and open new avenues for non-invasive microbiome monitoring. If this trial is successful, the findings could have significant implications for understanding host-microbiome interactions and developing novel diagnostic tools for various health conditions.

## Data Availability

The anonymized data will be available to everyone.

https://www.eur.nl/en/library/research-support/eur-data-repository

## Acknowledgement

This project was funded in part through a grant (KICH2.V4P.22.015) of the Dutch Organization for Scientific Research and the Dutch Cancer Foundation.

